# Impacts of people’s learning behavior in fighting the COVID-19 epidemic

**DOI:** 10.1101/2020.08.02.20166967

**Authors:** Baolian Cheng, Yi-Ming Wang

## Abstract

This work presents a mathematical model that captures time-dependent social-distancing effects and presents examples of the consequences of relaxing social-distancing restrictions in the fight against the novel coronavirus epidemic. Without social distancing, the spread of COVID-19 will grow exponentially, but social distancing and people’s learning behavior (isolating, staying at home, wearing face masks, washing hands, restricting the size and frequency of group gatherings, etc.) can significantly impede the epidemic spread, flatten the infection curve, and change the final outcome of the COVID-19 outbreak. Our results demonstrate that strict social distancing and people’s learning behavior can be effective in slowing the spread rate and significantly reducing the total number of infections, daily infection rate, peak of daily infections, and duration of the epidemic. Under strict social distancing, the rise and fall of infections would be nearly symmetric about the peak of of daily infections, and the epidemic spread would be essentially over within 60 days. Relaxing social distancing and people learning behaviors will significantly increase the total and daily numbers of infections and prolong the course of the outbreak. These results have immediate applications for the implementation of various social-distancing policies and general significance for ongoing outbreaks and similar infectious disease epidemics in the future (LA-UR 20-22877).

**Disclaimer:** This material is not final and is subject to be updated any time. Contact information.)

## 1. INTRODUCTION

The highly contagious and asymptomatic transmission nature of the novel coronavirus has led to the explosive spreading of the COVID-19 epidemic [1–4] and has drastically affected the world economy. Unlike past flu seasons (which required no social distancing because vaccines were available), slowing and controlling the COVID-19 pandemic requires immediate physical isolation, social distancing, and even community shelter-inplace orders. Without any intervention, the infection will follow a natural exponential growth path in time until most of the population is infected [5]. Social distancing plays a critical role in reducing the spread of the epidemic and flattens the infection curve. Many flu-based epidemic models [6-15] have been used to model the current pandemic, but because they do not take social distancing into account, these models have missed predicting the rate of spread and peak time of new infections. Recent studies [16-19] have attempted to address this issue. In this letter, we apply a novel epidemic mathematical model recently developed by Cheng and Wang [5] to quantify the impacts of social distancing and people’s learning behavior (isolating, wearing face masks, washing hands, avoiding gathering in groups, etc.). The model can be applied to a community of any size (country, state, county, or city) to predict the number of total infections, infection rate, time of peak new daily infections, and time to reach a plateau for cumulative infections (96% of the total infections of the epidemic). This study provides guidance for policy makers on when to reopen their community and economy.

## 2. IMPACTS OF SOCIAL DISTANCING

Cheng and Wang recently developed a mathematical model that describes the COVID-19 epidemic [5]. This model is based on the principle of supply and demand for the virus and takes into account social distancing and people’s learning behavior. The model provides analytic solutions for the trajectory of the epidemic spread weeks in advance, including the number of total infections, daily infection rate, time of peak new infections, and time to reach the plateau. The total infected population *P* (*t*) at a given time and the number of daily new infections in the model are described by functions,

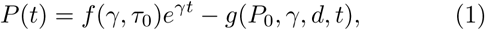

and

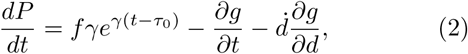

where *f* and *g* are functions of transmission rate (*γ*), social distancing (*d*), and the number of infected people (*P*_0_) at time *t*_0_. The social distancing level parameter *d* has values between *d*_*min*_ (no isolation or social distancing) and 1 (complete isolation, or infinite social distancing) depending on people’s learning behavior. The minimum value of *d* is determined by *d*_*min*_ ≡ *P*_0_*/*[(1 − *η*)*P*_*max*_], where *η* represents the fraction of the population who are naturally immune to the virus, and *P*_*max*_ is the total population of the community. The term 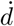 denotes the time derivative of *d*, which is important when social distancing is not constantly maintained over time. Clearly, increased social distancing with time 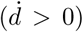 would reduce the daily infection rate. On the other hand, relaxing social distancing over time 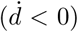 will increase the daily new infection rate. The parameters *γ* and *d* are calibrated to data from a given community at the time when social distancing was put into effect.

We extend the meaning of social distancing to include physical isolation, sheltering in place, staying at home, wearing face masks in public, washing hands, and restricting group-gathering size and frequency. If no social distancing is implemented, *d* = *d*_*min*_ ≈ 0, *g* = 0 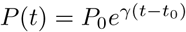 and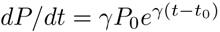, and both the total and daily infections grow exponentially with time until the number of susceptible individuals is depleted, or nearly all people, *P*_0_*/d*_*min*_ = (1 − *η*)*P*_*max*_, have been infected. But if all the infected people are clearly traced, identified, and completely isolated, *d* = 1, *P* (*t*) = *P*_0_ and *dP/dt* = 0, there would be no spread at all and, in turn, no epidemic.

In the absence of vaccines, people’s learning behavior is critical to controlling the spread of COVID-19. Given the community transmission rate *γ*, the behavior of people will, in principle, determine the outcome of the epidemic, which includes the total number of infections and deaths, time of peak new daily infections, and the time to reach a plateau in the total number of infections and deaths. To illustrate these effects, we take the transmission rate *γ* ≃ 0.17/day (or the effective reproduction number *R*_0_ ∼ 1.2 – 2.4), as the average transmission rate observed in the United States and assume *P*_0_ = 10000 at time *t*_0_, when the social distancing is put in place. We also assume the social-distancing level remains constant from time *t*_0_ through the epidemic. The total number of infections at a given time and the daily infection rate under various levels of social distancing (*d* = 0, 0.003, 0.005, 0.01, 0.02, 0.03) can be plotted as a function of time *t*, as shown in Fig. 1.

**FIG. 1.**
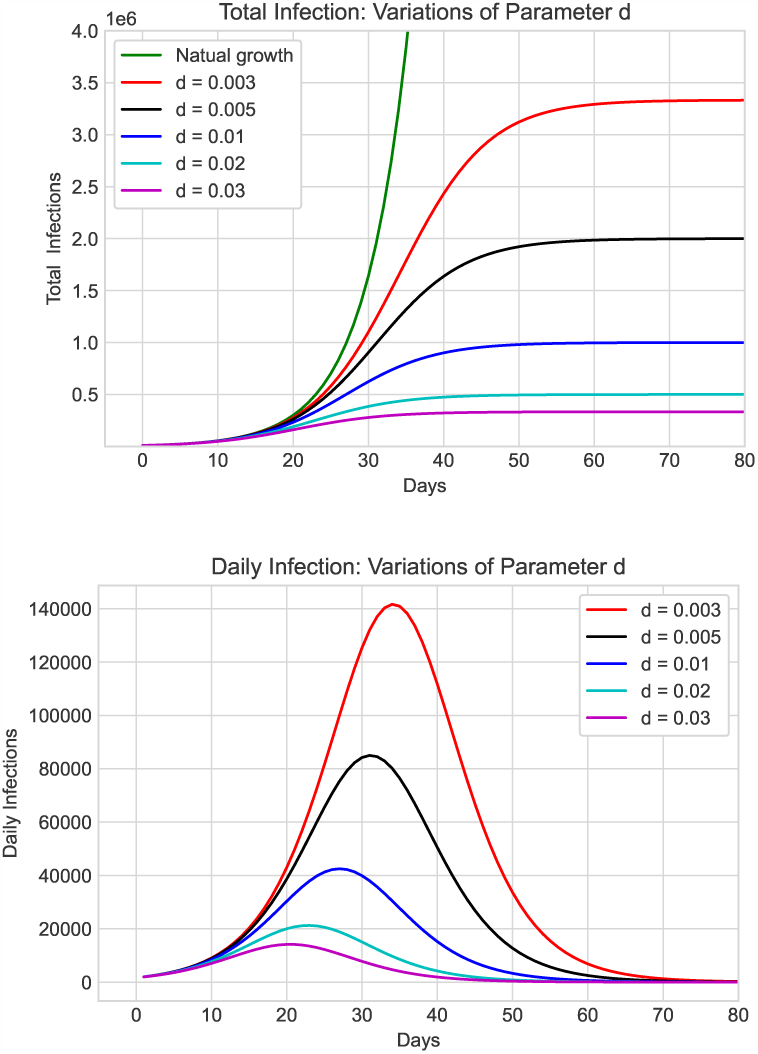
Total number of infections (top) and daily number of new cases (bottom) versus time as functions of the social distancing parameter *d*. The red, black, blue, cyan, and purple lines represent increasing social distancing at *d* = 0.003, 0.005, 0.01, 0.02, and 0.03, respectively. The green solid line in the top figure shows the total infections with no intervention, where *γ* = 0.17.

The infection data for the US [20, 23, 24] show that the social distancing level across the United States around March 22, 2020 ranged from 0.015 (New Jersey) to 0.065 (San Francisco) [5]. Our results show that the number of infections–both the total number over the course of the epidemic and the daily number of new infections–are very sensitive to people’s learning behavior and the level of social distancing achieved. Both quantities dramatically increase when social distancing is relaxed, and the date of the peak in daily infections is delayed. This result differs from other studies, which capture the delaying effect of social distancing on the peak number of infections but not the effects on the total and daily number of infections [16-19]. Our model clearly shows that reducing social distancing not only increases the length of time until life returns to “normal” but also places more lives at risk.

The above analysis assumes a constant level of social distancing, but people’s behavior can change during an epidemic. For example, after a period of sheltering in place, various pressures (psychological, economic, etc.) may encourage people to relax their social distancing behavior once the peak of daily infections has passed in their community. Hot weather may result in fewer people wearing face masks. Recent studies show that the combination of using face covers, keeping a physical distance between individuals, and washing hands is the most effective and cost-saving strategy in the battle against COVID-19 [25]. The asymptomatic transmission between two people can be reduced significantly from 100% if no one wears a face mask to ∼ 1.5% if everyone wears a face mask. In both South Korea and Japan, businesses were never shut down, but people did wear face masks all the time and in all places. Their infection rates peaked early (March 4 in South Korea and April 15 in Japan), and both countries have relatively few new infections.

Based on the above information, we can summarize the effect of relaxing social distancing requirements on people in two explicit mathematical expressions: (1) people reducing their wearing of face masks in public and (2) people reducing their distance of close contact and increasing the size and frequency of occasional group gatherings. We assume the social-distancing parameter changes with time as *d* = *d*_0_*e*^−*βt*^[1 + *k* sin(2*πt/T*)], where the parameter *β* represents a relaxation rate representing the percentage of people who stop wearing masks each day and *T* is a relaxation time representing a time period over which people relax their social behavior, for example, by gathering in groups with less than 6 feet of separation.

We illustrate the effect of changes in people’s behavior with an example using the average US spread rate *γ ∼* 0.17 in late March 2020 [5]. We consider four scenarios: (1) the level of social distancing remains constant throughout the epidemic at the level of the week of March 23, 2020, *d* = *d*_0_ ≃ 0.04, *β* = *k* = 0; (2) the level of social distance drops because a large portion of the population does not wear face mask in public and starts gathering in groups every 2 weeks, corresponding to *d*_0_ = 0.05 and a relaxation rate *β* ≃ 1.35%/day, *k* = 0.002, and *T* = 11 days; (3) the level of social distance drops because a modest portion of people do not wear face masks in public spaces and they start gathering every 4 weeks, *d*_0_ = 0.048, *β* = 0.9%/day, *k* = 0.0329, and *T* = 24 days; and (4) the level of social distance drops because a very small fraction of people do not wear face masks in public and they start gathering weekly, *d*_0_ = 0.044, *β* = 0.35%/day, *k* = 0.002, and *T* = 7 days. Figure 3 shows the effects of these changes in social distancing and people’s learning behavior.

**FIG. 2.**
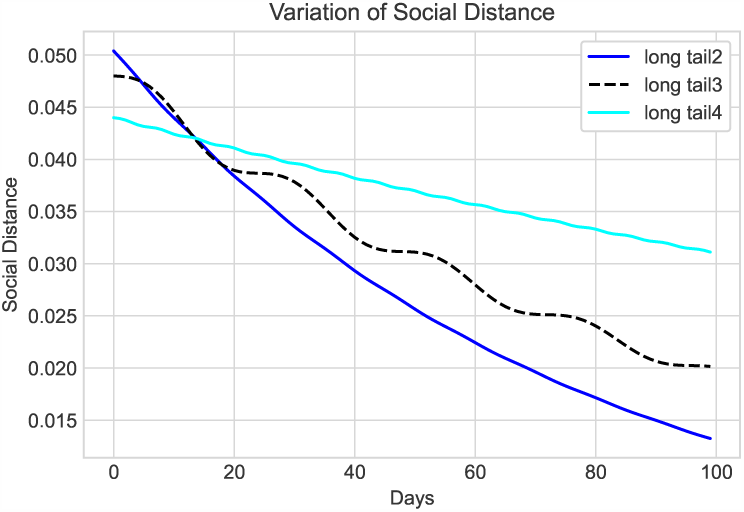
The effect of social distancing parameter *d* changing with time. The blue line represents the fast decline of social distancing with a time constant of 11 days (high proportion of people with no face mask). The black dashed line represents slightly less relaxing of social distancing over a longer relaxation period of 24 days. The cyan line denotes a slightly reduced social distancing but with a relaxation period of 7 days.

**FIG. 3.**
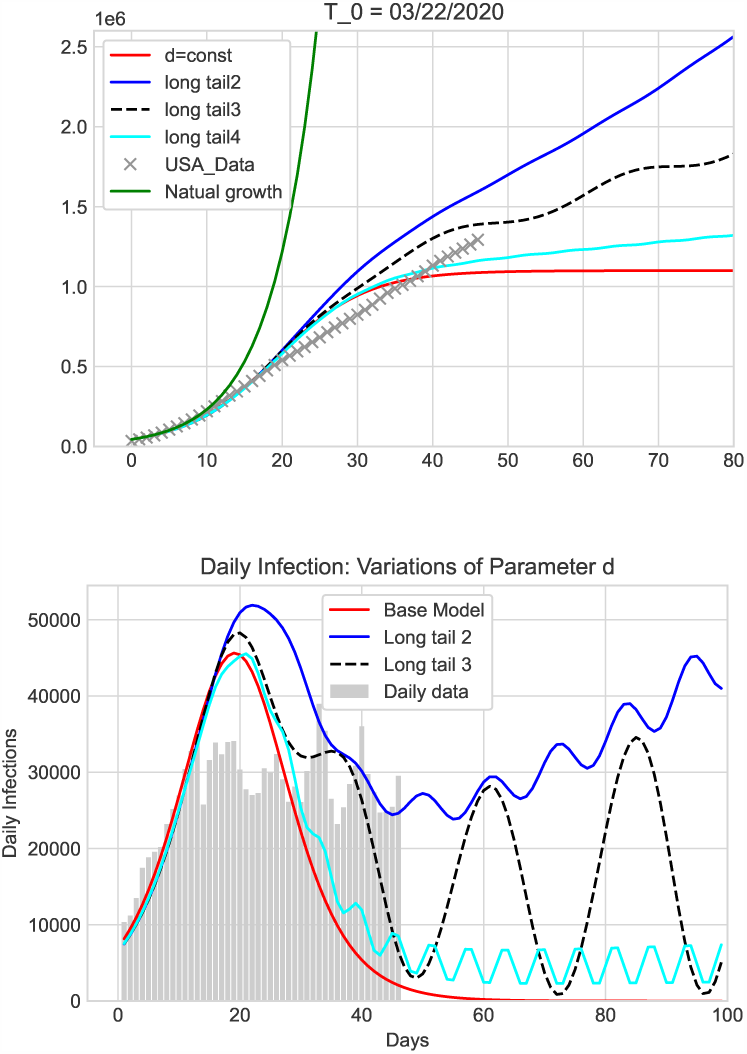
Total number of infections (top) and number of daily new cases (bottom) changing over time under four socialdistancing scenarios. The red line represents the spread trajectory under constant social distancing at the level when restrictions were put in place. The blue line corresponds to social distancing decreasing at a relaxation rate *β* = 1.35%/day and relaxation time of 11 days. The black dashed line corresponds to 0.9%/day and 24 days. The cyan line corresponds to 0.35%/day and *T* = 7 days. All cases assume a community spread rate of *γ* = 0.17.

Figure 3 shows that the outcome and spread trajectory of COVID-19 is very sensitive to the level of social distancing and people’s behavior. The total number of infections increases with the number of people not wearing face masks in public and decreases with the number of days of people maintain social distancing. The more people wear face masks in public, the faster the epidemic ends and the lower the number of infections. With fewer people wearing face masks, the epidemic lasts much longer, and the number of daily infections oscillates for a long time (similar to the blue and cyan lines shown in Fig. 3). The epidemic spread can even grow into a second wave (as shown by the black dashed line in Fig. 3). When social distance is maintained at a constant level of first implementation, the number of new cases decreases symmetrically about the peak of daily infections, and the epidemic is over within 60 days from *t*_0_. This result agrees with the epidemic data reported from South Korea [21, 22], Japan [26] and some other countries.

## 3. APPLICATIONS

Applying our analysis to a number of pandemic centers worldwide and comparing our results with epidemic data reported by the Johns Hopkins University [20] and www.worldometer.info, we find that the data for countries like Japan [26], South Korea [21], France [27], Germany [28], Norway [29], etc. are consistent with a steady social distancing maintained throughout the epidemic (e.g., March 20 to June 7), while the epidemic data for the United States [23], United Kingdom [30], and Sweden [31] are consistent our hypothetical scenario (2), in which the number of people wearing face masks in public drops significantly over time. Figure 4 shows the epidemic data from France and Sweden. The epidemic trajectory of Iran [32] matches scenario (3), reflecting with a longer relaxation time, while Singapore [37] and many European countries, such as Spain [37], Italy [34], Denmark [36], Finland [35], etc., are consistent with scenario (4). Figure 5 shows the data from Iran, Singapore, and Spain.

**FIG. 4.**
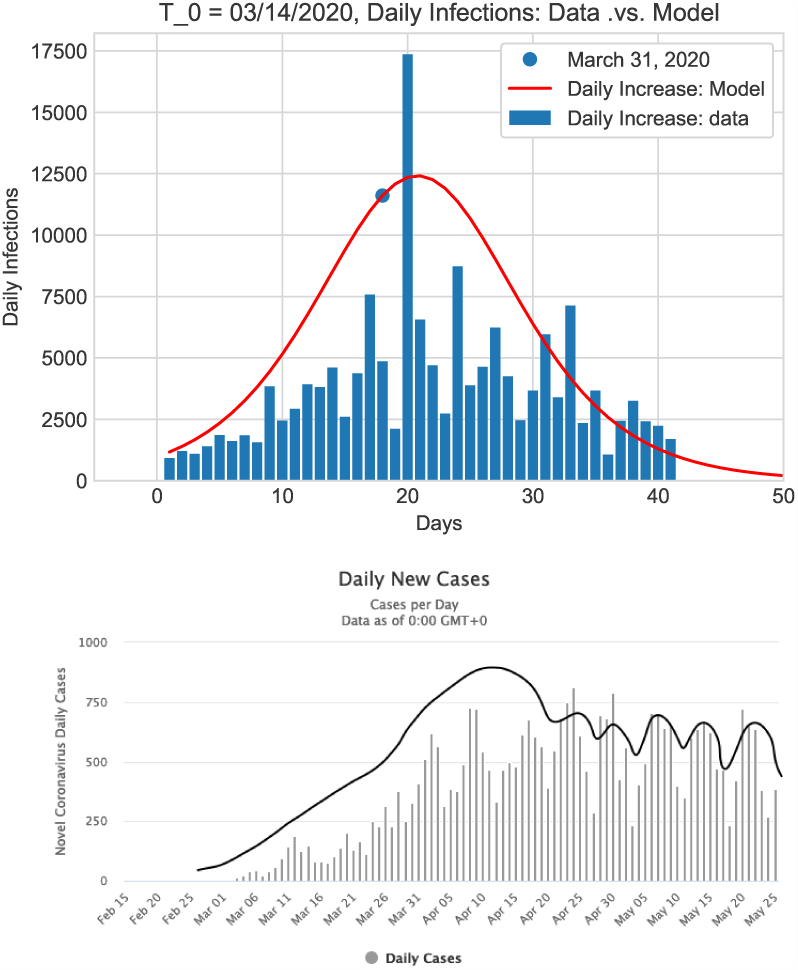
Number of daily new infections as a function of time for France (top) and Sweden (bottom). The red line in the top plot is the daily infections from our model with constant social distancing *d* from the week of March 20, 2020. The line in the bottom plot is the envelope of the data (not from our model). When compared to the blue curve in Fig. 3, the shape of data indicates periodically relaxed social distancing during the epidemic and a notable relaxation rate of people wearing face masks in public areas.

**FIG. 5.**
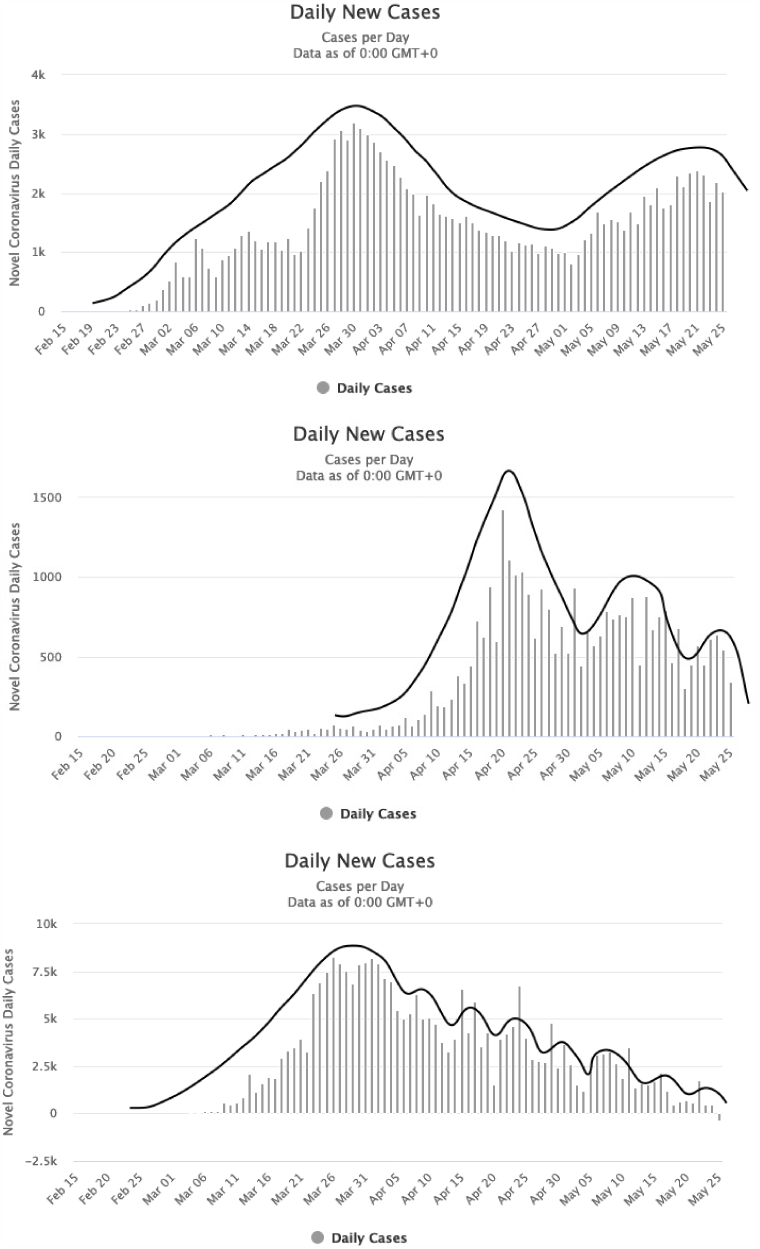
Number of daily new infections as a function of time for Iran (top), Singapore (middle), and Spain (bottom). The line in each figure is the envelope of the data (not from our model). The epidemic trajectory in Iran suggests that a longer relaxation time is occurring, while the trajectories for Singapore and Spain suggest that most people are wearing masks but the relaxation time is shorter, consistent with more frequent group gatherings.

Our analysis shows that social distancing and changes in people’s behavior can significantly affect the cumulative and daily number of infections and, accordingly, the number of deaths over the course of a pandemic. Good social distancing includes (a) wearing face masks in public spaces, (b) not gathering in groups over a certain size, (c) washing hands frequently, and (d) always keeping minimal 6 feet from the others. When people follow these practices, communities will be able to reopen their economies sooner and minimize the number of infections in the absence of a vaccine.

## 4. CONCLUSION

Strict social distancing and significant changes in people’s learning behavior can play significant roles in reducing the spread of the novel coronavirus during this pandemic. Without social distancing, the spread of COVID-19 would grow exponentially with time until most people (∼70%) are infected. Social distancing and changes in people’s behavior (isolation, wearing masks in public spaces, restrictions on group gathering size, washing hands, etc) will significantly reduce the virus spread rate and, in turn, dominate the final outcome of the COVID-19 pandemic. We showed the dependence and sensitivity of the number of total and daily infections on social distancing under four different scenarios (1) maintaining constant social distancing; (2) a high relaxation rate of people wearing face masks in public spaces and gathering in groups every 2 weeks; (3) a modest relaxation of people wearing face masks in public spaces and gathering in groups every 4 weeks. and (4) a much smaller relaxation rate of people wearing face masks in public spaces and gathering in groups every week. Our results show that strict social distancing and changes in people’s behavior will not only slow the virus spread rate but also significantly reduce the number of total infections, daily infection rates, peak of daily infections, and duration of the epidemic as a whole. Under strict social distancing, the rise and fall of the epidemic spread are nearly symmetric around the peak of daily infections, and the total duration of the epidemic would be less than 60 days. If everyone wears a face mask in public areas, there would not be a need to shut down the economy even if there was a second wave of COVID-19. Relaxed social distancing will result in many more infections and deaths. These research results have immediate applications in the implementation of various social distancing policies and general significance for ongoing outbreaks and similar infectious disease epidemic in the future.

## Data Availability

The data that support the findings of this study are
openly available. Code used will be made available as soon as possible.

## ACKNOWLEGEMENTS

The authors are grateful to Dr. Arick Wang for many valuable discussions, Dr. Paul A. Bradley for valuable comments, and Hang Zhou for the data of US states and valuable discussions. The authors are grateful to C. S. Carmer for editing this article. B.C. was supported under the auspices of the U.S. Department of Energy by the Los Alamos National Laboratory under Contract No. 89233218CNA000001.

## Notes

### Competing Interest Statement

The authors have declared no competing interest.

